# Age reporting in the Brazilian COVID-19 vaccination database: What can we learn from it?

**DOI:** 10.1101/2021.06.27.21259575

**Authors:** Cássio M Turra, Fernando Fernandes, Julia Calazans, Marília R. Nepomuceno

## Abstract

Age is a key variable for sciences and public planning. The demographic consequences of not measuring age correctly are manifold, including errors in mortality rates and population estimates, particularly at older ages. It also affects public programs because target populations depend on reliable population age distributions. In Brazil, the start of the vaccination campaign against COVID-19 marked the collection of new administrative data. Every citizen must be registered and need to show an identity document to get vaccinated. The requirement of proof-of-age documentation provides a unique opportunity for measuring the elderly population using a different database. This article examines the reliability of age distributions of men and women 80 years and older. We calculate various demographic indicators using data from the vaccination registration system and compare them to those from the target population estimates of the National Vaccination Plan, censuses, and population projections for Brazil and countries with high-quality population data. We show that requiring proof-of-age, such as in the vaccination records, increases data quality, mainly through the reduction of age heaping and age exaggeration. However, I.D. cards cannot fully solve wrong birth dates that result from weak historical registration systems. Thus, one should be careful when using estimates of the old age population living in some of the Brazilian regions, particularly the North, Northeast, and Center-West. Also, our analysis reveals a mismatch between the projected population by age, sex, and region, which guided the vaccination plan, and the number of vaccinated at ages 80 and older. The methodology developed to adjust the mortality rates used in the demographic projections is probably the main factor behind the disparities found.

## Introduction

Age is a key variable for sciences and public planning. The demographic consequences of not measuring age correctly are manifold, including errors in mortality rates and population estimates, particularly at older ages (Preston et al. 1999). It also affects public programs because target populations depend on reliable population age distributions.

Age misreporting varies across data sources, countries, population subgroups, and periods (Coale and Kisker 1986; Elo and Preston 1994, Dechter and Preston 1991, Nepomuceno and Turra 2020, Gomes and Turra 2009). Research has associated it with different factors, including delayed birth registration, which precludes individuals from knowing their actual age, response errors from memory recall problems, proxy reporting, and low numeracy and literacy. Most household surveys and administrative records worldwide now include a question about the birth date instead of age in completed years, improving data quality, and mitigating age heaping. Still, in places with insufficient quality birth registration data, recalling the actual birth date may remain challenging, particularly among the elderly of low socioeconomic status (SES).

In Brazil, data are not free of age misreporting. Turra (2012) has claimed that the pattern of mortality at older ages in Brazil, characterized by rates relatively lower than in high-income countries, reflects age exaggeration. Recent findings have corroborated his argument (Nepomuceno and Turra 2020; Di Lego, Turra, and Cesar 2017). Also, the excess of centenarians in official Brazilian data is consistent with age misreporting due to delayed birth registration during the nineteenth and twentieth centuries (Nepomuceno and Turra 2020; Gomes and Turra 2014). Studies on adult mortality for Latin America suggest that the quality of age reporting is probably not limited to the oldest-old and may affect population estimates and deaths at younger ages (Dechter and Preston 1991; Rosenwaike and Preston 1984; Grushka 1996; Glei et al. 2021). Therefore, any new database that keeps consistent birth date records can significantly improve social programs, population estimates, and projections.

During a global public health crisis, such as the COVID-19 pandemic, the demand for high-quality information increases, and there are opportunities for building new databases. In Brazil, the start of the vaccination campaign marked the collection of new administrative data. The National Plan for Operationalization of the Vaccine against COVID-19, implemented by the Federal Government, defined different priority groups, including individuals 60 and older (Brasil 2021a). Every citizen must be registered and need to show an identity document to get vaccinated. The requirement of proof-of-age documentation provides a unique opportunity for measuring the elderly population using a different database.

Identity documents cannot solve the wrong ages from delayed birth registration. Still, they can mitigate age misreporting when response errors originate from memory recall problems, proxy reporting, or low numeracy and literacy. Therefore, we expect more accurate age distributions for the vaccinated than in the official population census and population projections grounded on data sources that are more vulnerable to reporting errors. We hypothesize that older population groups are overestimated in the National Vaccination Plan due to the known age exaggeration at these age groups in Brazil (Turra 2012; Nepomuceno and Turra 2020). If this is the case, vaccination coverage rates by age may be misleading and should be taken cautiously.

This article examines the reliability of age distributions of men and women 80 years and older. To achieve our goal, we calculate different demographic indicators using data from the vaccination registration system and compare them to those from the target population estimates of the National Vaccination Plan, censuses, and population projections for Brazil and countries with high-quality population data. Further, we look at results by Brazilian regions since age reporting is associated with socioeconomic conditions. Our analyses make two critical contributions to the demographic and public health literature. First, it offers new critically assessed estimates of the Brazilian oldest old population based on the vaccination registration system. Second, it uncovers potential problems in the estimated number of vaccines needed to cover individuals that should be age-prioritized, compromising the efficiency and speed of the vaccination campaign.

## Materials

### Vaccination Data

We drew data from the Brazilian Ministry of Health open microdata registers for the national COVID-19 vaccination campaign (Brasil 2021b). We used vaccination registers recorded from the official beginning of the campaign (January 18, 2021) until June 05, 2021. The database comprises anonymized information on both first and second doses of any COVID-19 vaccine. It also includes data on age, date of birth, sex, race, type of dose (first or second), vaccination group (e.g., health worker, age group), city/state of vaccination and residence, and vaccine name and developer.

We considered only the population vaccinated with the first dose of any COVID-19 vaccine for Brazil and its regions, by sex and single ages from 80 to 110 and older. Limiting the age range is necessary since the vaccination program is still ongoing. The initial database comprises 66,821,493 records, of which 648,248 are either duplicated (639,956), triplicated (8,286), or quadruplicated (6). Therefore, after cleaning for identical records, the database consisted of 66,173,245 cases (99.03% of the initial database). From this total, we excluded records that had missing dose (2,993; 0.005%), missing sex (70; 0.0001%), and vaccination date before January 18, 2021 (7,407; 0.011%). There are also 6,311 (0.009%) individuals for whom the date of birth is December 30, 1899 (121 years old). Among them, 5,540 are healthcare workers. Thus, we believe this is the system default for unknown birth date and excluded the 5,540 (0.008%) cases from our database. Another 346,501 (0.52%) cases have a missing state of residence. We fixed these cases by obtaining the state of residence from the state of vaccination which has no missing values. Finally, we filtered 20,678,703 (31.25%) cases that are related to the second dose.^3^ Although old-age individuals may have died between the beginning of the vaccination campaign and June 06, 2021, we cannot identify who they are. Therefore, we ignored any deaths and estimated the vaccinated population as of May 15, 2021, the last date with available information regarding COVID-19 excess mortality for 2021 (CONASS 2021). Our final database consisted of 4,375,174 individuals 80 years and older, 2,711,186 females, and 1,663,988 males.

### Population Estimates

We compared the vaccinated population with four other estimates for the Brazilian older adults, including the target population defined by the Brazilian Ministry of Health (MoH) (Brasil 2021a), Brazilian Census from 1960 to 2010 (Minnesota Population Center 2020), and population projections prepared by the Brazilian Institute of Geography and Statistics (IBGE 2018) and the United Nations (United Nations 2019). The target population, defined by the MoH in the national plan, is the IBGE population projection for July 1, 2020. It presents estimates for both sexes only and by five-year age groups from 60 to 90 and older. We did not make any adjustments to the target population since it is the official population distribution for the vaccination campaign. The IBGE original projections by five-year age groups and sex are also limited to the open age group 90+ but include Brazilian regions and states. In contrast, the UN projections by five-year age groups and sex are restricted to Brazil’s total geographical area but have age groups up to 100+.

To make the population projections (IBGE and the UN) comparable to the vaccinated population, we used exponential interpolation by 5-year age groups and sex to adjust the figures to May 15, 2021. Additionally, we subtracted from the interpolated distributions the excess of deaths from January 1 of 2020 to May 15, 2021, due to the COVID-19 pandemic. To do this, we applied the proportion of excess of deaths by sex, year (2020 and 2021), and region of residence for ages older than 60, estimated by CONASS (2021), to the number of deaths implicit in each population projection (IBGE and the UN). We also compared the Brazilian vaccination records against data from countries known by high-quality population enumerations at old ages. Our group includes Japan (2005, 2010, and 2015), Sweden (from 1992 to 2019), and Switzerland (from 2002 to 2018) by age and sex. We used raw data from the *Human Mortality Database* (2021) and selected the years with population enumerations that include ages 110 and older.^4^

We tabulated population numbers by age groups, sex, and regions. Then, we calculated the proportionate age distributions, age ratios 100+/80+ and 90+/80+, and compared them across the different data sources. We also displayed the age ratios against mortality levels measured by life expectancy at age 50 (IBGE 2018; Human Mortality Database 2021). Further, we estimated the blended Myers index (Myers 1954) to measure levels of age heaping for ages 80 to 99 in the vaccination records and Brazilian censuses. Finally, we estimated vaccination coverage ratios using the adjusted-to-COVID-19 IBGE population projections as the denominators.

## Results

Between May 30, 2021 and June 05, 2021 - the last seven days before our database cutoff date - daily growth rates for the vaccinated population ages 80 and older have been low in all Brazilian regions. It ranged from 0.001% to 1,002%. Despite the minor variations over time, we do not compare absolute differences because they involve more uncertainty than relative measures.

Table 1 shows that the ratios 90+/80+ and 100+/80+ vary across data sources. First, according to the MoH and IBGE, the ratio 90+/80+ for both sexes would be 0.201 and 0.180, respectively, while the UN projects a younger age distribution (0.161). Vaccination records present an intermediate scenario (0.174), which is younger than the MoH and IBGE estimates but older than the UN. Although vaccination records’ 90+/80+ ratio is only 8% higher than the UN’s, its 100+/80+ ratio is almost twice as high.

**Table 1.** Population age distributions at ages 80 and older by sex. Brazil. Selected data sources.

| Open-ended age groups<br>ratio and absolute<br>values by sex | Vaccine records, as<br>of June 5th, 2021, age<br>adjusted to May 15th,<br>2021 | Population Estimates |  |  |  |  |
| --- | --- | --- | --- | --- | --- | --- |
|  |  | Ministry of<br>Health<br>(MoH),<br>Target<br>Estimates | IBGE<br>rev. 2018 |  | United Nations Population<br>Division<br>rev. 2019 |  |
|  |  |  | Original<br>Projections as<br>of May 15th,<br>2021 | Adjusted to<br>excess deaths | Original<br>Projections as<br>of May 15th,<br>2021 | Adjusted to<br>excess deaths |
| *Ratio 90+/80+* |  |  |  |  |  |  |
| Both sexes | 0.174 | 0.201 | 0.185 | 0.180 | 0.166 | 0.161 |
| Women | 0.189 | - | 0.202 | 0.197 | 0.183 | 0.178 |
| Men | 0.149 | - | 0.158 | 0.152 | 0.137 | 0.132 |
| *Ratio 100+/80+* |  |  |  |  |  |  |
| Both sexes | 0.009 | - | - | - | 0.005 | 0.005 |
| Women | 0.011 | - | - | - | 0.006 | 0.006 |
| Men | 0.007 | - | - | - | 0.004 | 0.003 |

Source: Prepared by the authors, based on Brasil. 2021. "Campanha Nacional de Vacinação contra Covid-19 - Registros de Vacinação COVID-19 - Open Data." openDataSUS. 2021; Brasil. 2021; Plano Nacional de Operacionalização da Vacinação Contra a Covid-19. 6th ed. Brasília, DF; Conselho Nacional de Secretários de Saúde - CONASS. 2021. "Painel de Análise do Excesso de Mortalidade Por Causas Naturais No Brasil." 2021; IBGE. 2018. Projeções da População: Brasil e Unidades da Federação - Revisão 2018. 2nd ed. Séries Relatórios Metodológicos, volume 40. Rio de Janeiro: IBGE, Coordenação de População e Indicadores Sociais; United Nations. 2019. World Population Prospects 2019: Online Edition. New York: United Nations, Department of Economic and Social Affairs, Population Division.

To further assess the quality of data on age, we compared Myer’s summary preference index (ages 80-99) and the proportion of centenarians (100+/80+) for the vaccination records with those for the Brazilian censuses (1960 to 2010). Table 2 shows that the degree of age heaping lessened to about a quarter between the 1960 and 2010 censuses. Yet, the reduction was not monotonic. The index increased between the 2000 and the 2010 census for the Northeast, Center-West, and Southeast regions. In 2021, the vaccination records showed 30 to 52% lower indexes than the 2010 census for all regions. Considering digit preference was already down in 2010, the improvement was substantial.

**Table 2:** Myer’s summary preference index (ages 80-99) and the proportion of centenarians (100+/80+). Brazil and its regions. Vaccination records (2021) and Census Data (1960-2010)

| Data/Year | Regions/Measures |  |  |  |  |  |  |  |  |  |
| --- | --- | --- | --- | --- | --- | --- | --- | --- | --- | --- |
|  | North |  | Northeast |  | Center-West |  | Southeast |  | South |  |
|  | Myers's Index | 100+/80+ (%) | Myers's Index | 100+/80+ (%) | Myers's Index | 100+/80+ (%) | Myers's Index | 100+/80+ (%) | Myers's Index | 100+/80+ (%) |
| Census Data |  |  |  |  |  |  |  |  |  |  |
| 1960 | NA | NA | 24.3 | 4.21 | 24.6 | 5.95 | 15.9 | 4.21 | 11.8 | 3.73 |
| 1970 | 15.7 | NA | 19.1 | NA | 17.5 | NA | 11.3 | NA | 9.4 | NA |
| 1980 | 8.8 | 3.24 | 10.1 | 2.48 | 8.4 | 4.21 | 6.7 | 1.65 | 6.4 | 2.19 |
| 1991 | 5.0 | 2.21 | 7.0 | 1.62 | 4.9 | 1.93 | 4.8 | 1.29 | 3.1 | 1.38 |
| 2000 | 4.9 | 2.86 | 4.2 | 1.70 | 3.6 | 2.45 | 4.2 | 1.56 | 5.2 | 1.32 |
| 2010 | 4.3 | 2.07 | 6.2 | 1.66 | 5.0 | 1.82 | 4.5 | 1.15 | 4.1 | 1.11 |
| Vaccination Records |  |  |  |  |  |  |  |  |  |  |
| 2021 | 2.6 | 1.69 | 3.2 | 1.62 | 3.6 | 0.84 | 2.2 | 0.64 | 2.7 | 0.51 |

Source: Prepared by the authors, based on Brasil. 2021. “Campanha Nacional de Vacinação contra Covid-19 - Registros de Vacinação COVID-19 - Open Data.” openDataSUS. 2021; Minnesota Population Center. 2020. “Integrated Public Use Microdata Series, International: Version 7.3.” Minneapolis, MN: IPUMS. https://doi.org/10.18128/D020.V7.3.

The proportion of centenarians also reduced substantially (about 70%) over the 50 years, despite survival gains, indicating census data improvement. The exception is the year 2000 when the proportion increased for all regions, except the South. In 2021, the vaccination records showed further declines in the 100+/80+ ratio, particularly in the South and Center-West (54%) and the Southeast (45%). Unfortunately, since the IBGE postponed the 2020 census, we do not know to what extent quality improvement in the vaccination records reflects a historical trend, proof of age requirement, or a combination of both. Still, increases in age reliability with the 2021 vaccination records are more significant than any census improvements after 1991.

Variations in the indicators across Brazilian regions reflect historical patterns of socioeconomic development. Age misreporting is usually a more significant problem where vital registration systems have been weaker. In Brazil, data errors used to be an issue everywhere, but they were more important in the Northern and Central regions (North, Northeast, and Center-West states). Geographic distances, register costs, larger shares of rural and low SES populations explain the regional pattern of delayed birth registration during the twentieth century (Hakkert 1996). Therefore, the regional patterns of age exaggeration (100+/80+) and age heaping in Table 2 are not surprising. Figure 1 examines this finding in more detail by displaying 90+/80+ ratios against 100+/80+ ratios and e_50_ for the five Brazilian regions and Japan, Sweden, and Switzerland. Both nonagenarians and centenarians represent much larger shares in the North and Northeast regions. The exaggeration is more pronounced among men. For example, conditional on e_50_, the proportion of nonagenarians in the Northeast is about 54% (women) and 110% (men) higher than expected, giving the experience of the other countries. Only the Brazilian South (men and women) and Southeast (women) are within or close to the international high-quality-data-zone. Although historical variations in fertility and migration contribute to determining age distributions of adults, they likely play only a minor role at older ages.

**Figure 1.**
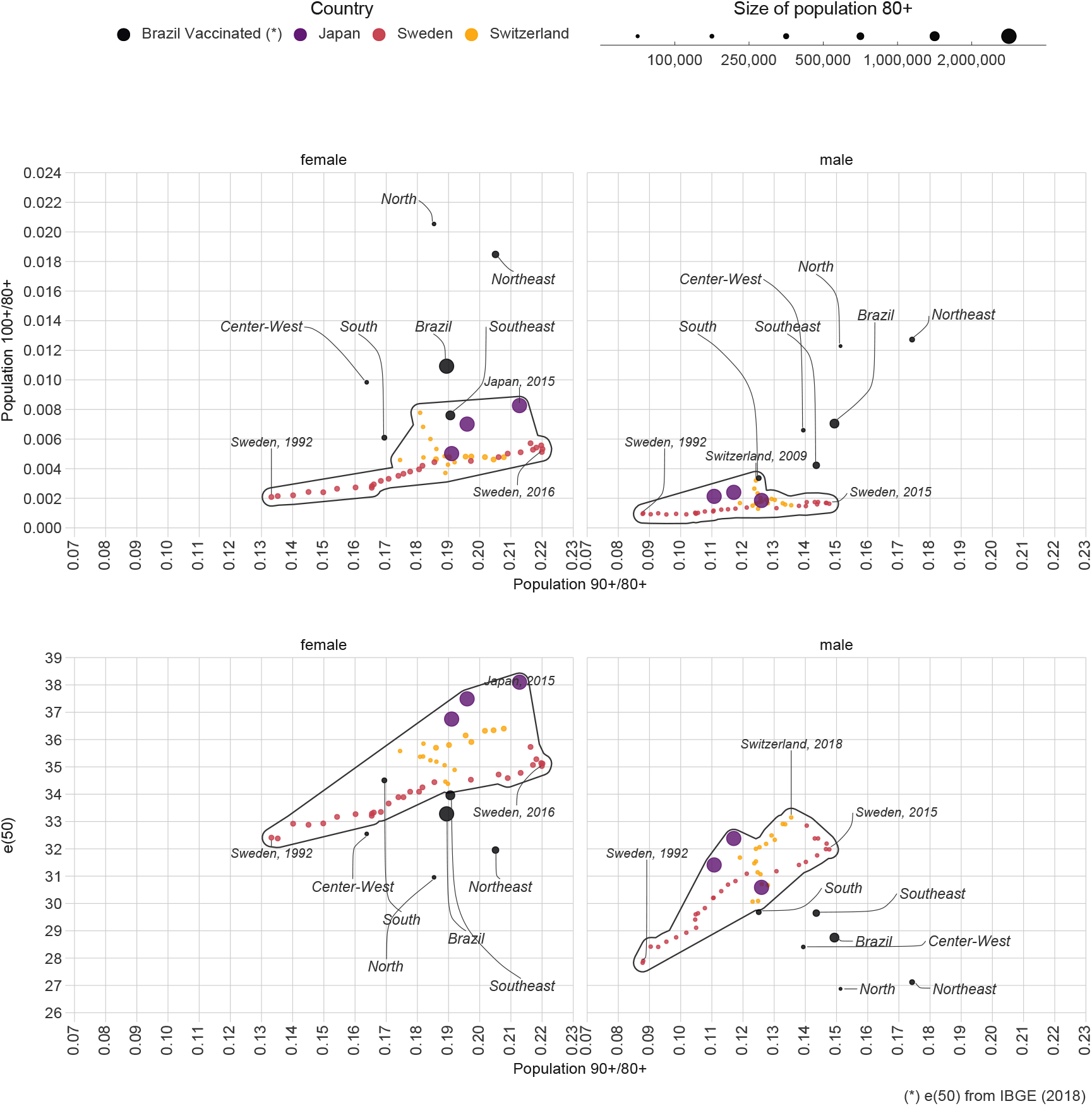
Proportion of individuals 90 and older (90+/80+) by the proportion of centenarians (100+/80+) and life expectancy at age 50. Men and Women. Brazil and its regions (vaccination records) and selected countries Source: Prepared by the authors, based on Brasil. 2021. “Campanha Nacional de Vacinação contra Covid-19 - Registros de Vacinação COVID-19 - Open Data.” openDataSUS. 2021; IBGE. 2018. Projeções da População: Brasil e Unidades da Federação - Revisão 2018. 2nd ed. Séries Relatórios Metodológicos, volume 40. Rio de Janeiro: IBGE, Coordenação de População e Indicadores Sociais; Human Mortality Database. 2021. University of California, Berkeley (USA) and Max Planck Institute for Demographic Research (Germany).

So far, we have shown that data quality on age is better in vaccination records than IBGE census and estimates. However, proof of age did not prevent vaccination records from the historical and regional patterns of errors. One obvious question is how IBGE deals with these issues since yearly demographic estimates by municipalities, states, and regions are necessary to inform public policies. The number projected in the old-age groups depends on the age distributions of mortality rates and populations at baseline. Figure 2 shows the proportion vaccinated for each region by dividing vaccination records and IBGE projections adjusted to excess deaths due to the COVID-19 pandemic. The regional-age-sex pattern of differences emerges once more: ratios are higher than one in the North, Northeast, and Center-West regions and lower than one in the South and Southeast regions. Also, the discrepancies between the two data sources are more pronounced at ages 90 and older and among men, who have proved to be more susceptible to age exaggeration in Brazil. For example, in the Northeast, the ratio is equal to 1.30 (men) and 1.18 (women) at ages 90 and older, and 1.15 (men) and 1.08 (women) in the age group 85-89. The corresponding ratios are 0.78 and 0.92 (men) and 0.79 and 0.95 (women) in the South.

**Figure 2.**
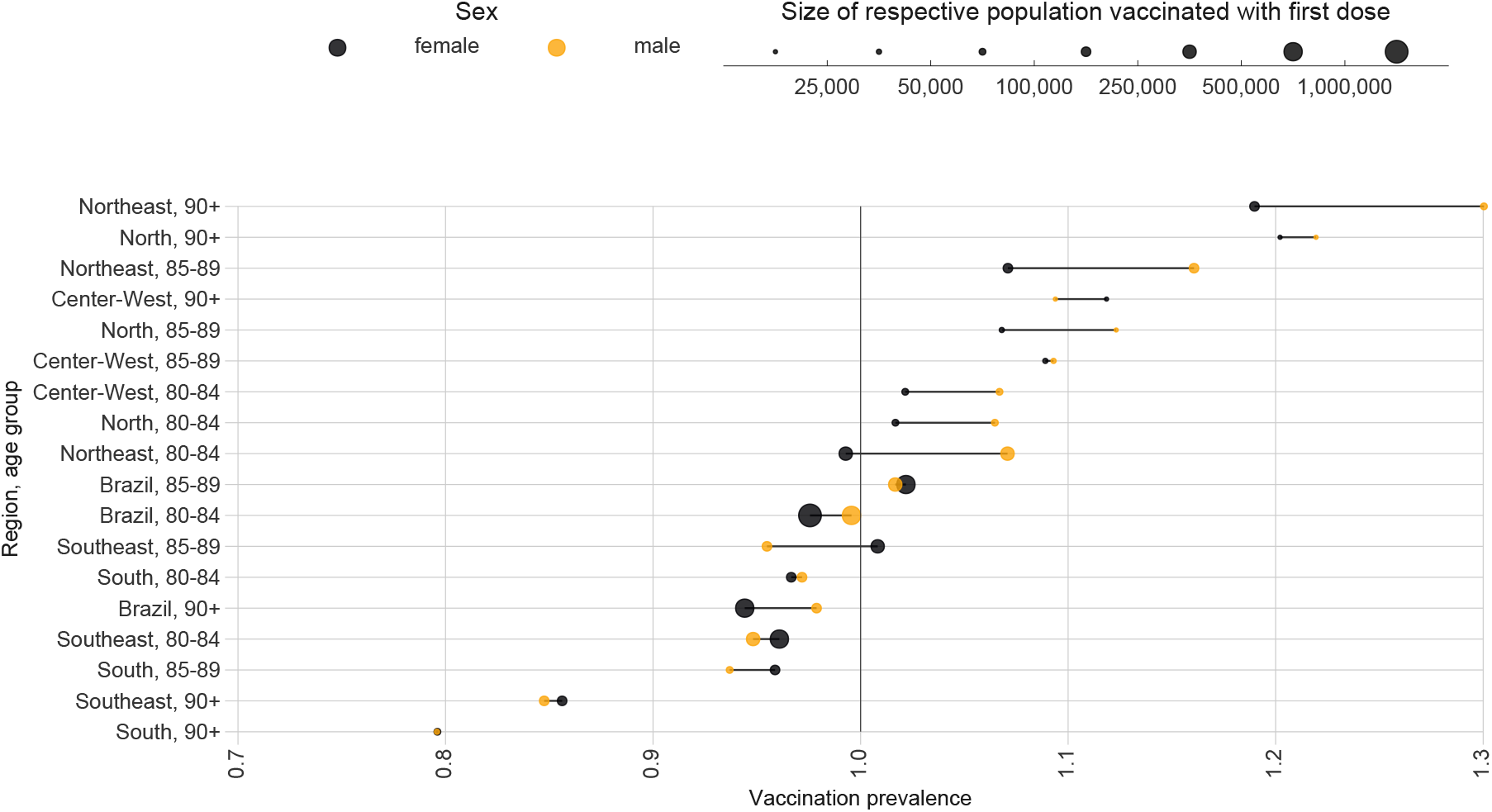
Vaccination coverage (ratio of vaccinated and projected populations) by age group and sex. Brazil and its regions, 2021 Source: Prepared by the authors, based on Brasil. 2021. “Campanha Nacional de Vacinação contra Covid-19 - Registros de Vacinação COVID-19 - Open Data.” openDataSUS. 2021; Brasil. 2021; Conselho Nacional de Secretários de Saúde - CONASS. 2021. “Painel de Análise Do Excesso de Mortalidade Por Causas Naturais No Brasil.” 2021; IBGE. 2018. Projeções Da População: Brasil e Unidades da Federação - Revisão 2018. 2nd ed. Séries Relatórios Metodológicos, volume 40. Rio de Janeiro: IBGE, Coordenação de População e Indicadores Sociais.

Why are the age distributions relatively older for the vaccinated than IBGE projections in the Northern states and vice-versa in the Southern states? One compelling hypothesis is the way IBGE (2013a) estimated mortality rates by region used in the projections. It first corrected the underreporting of child and adult deaths. It then proportionally reduced the number of deaths by age and sex in all regions, except for the North, Northeast, and Center-West (IBGE 2013, 24), to ensure the sum of corrected deaths for all regions meets the original number of deaths for Brazil as a whole. Increasing the number of deaths in the Northern and Central states relative to the others tries to adjust the unexpected lower adult mortality rates in these regions, whose data are more susceptible to coverage and content errors. However, the adjustment was made at the expense of reducing adult mortality in the South and Southeast. IBGE does not treat age misreporting issues explicitly.

As shown in this article, birth dates are wrong for a fraction of the elderly and cannot be fixed even with I.D. cards, particularly in the Northern regions. Therefore, one unexpected consequence of IBGE’s methodology is a projected population that does not match the observed regional distribution of the oldest-old in Brazil.

## Discussion

In agreement with our initial hypotheses, Brazilian data are not free of errors, which preclude us from knowing the exact number of individuals at older ages. Our analysis showed improvements in the age distribution of the oldest-old across the censuses. Also, requiring proof-of-age, such as in the vaccination records, increases data quality, mainly through the reduction of age heaping and age exaggeration. Yet, I.D. cards cannot fully solve wrong birth dates that result from weak historical registration systems. Thus, we cannot have confidence in estimates of the oldest-old for all regions of Brazil. Uncertainty is likely to increase for smaller areas such as states and municipalities.

As vital registration improves for the new birth cohorts, one can expect age reporting to become intrinsically more accurate in Brazil. Still, it will take some time to get precise age distributions at advanced ages. In the absence of high-quality vital records, historical census data have helped demographers identify the correct age of adults by tracking events and reports from the first years of life (Preston et al. 1996). Unfortunately, historical census microdata are missing for Brazil. The remaining option is to use statistical and demographic methods to adjust the current population and death counts by age (Palloni et al. 2021, Carrier and Farrag, 1959, Siegel and Swanson 2004). As aforementioned, the IBGE has estimated adjusted mortality rates for Brazilian regions, but it makes no explicit reference to age misreporting issues at older ages. It is unclear whether the current methodology is the best since the actual mortality patterns are unknown, and data errors may confound with other factors such as mortality selectivity. Also, the proposed adjustments resulted in transferring deaths across the regions. Whereas trying to adjust the mortality functions for coverage and content errors has enormous value and needs more research in Brazil, it resulted in population projections that do not match the actual distribution of individuals at older ages. Concerning public policies directed to the elderly, such as health-related campaigns, using the current population projections can be a problem since what counts for the policies is the citizens’ official birth dates.

Our article shows that any new database that keeps birth date records can be precious for population estimates in the Brazilian context. Systematic comparisons across old and new databases offer fresh clues about differences with the official figures. On the other hand, these comparisons can provide a critical assessment of the quality of the new databases. It is a two-way street in terms of reducing our ignorance about age and other demographic distributions. However, our analysis is not free of limitations. As with any database, vaccination records may contain both content and coverage errors. Concerning the potential coverage errors, some individuals ages 80 and older may have refused to vaccinate. Yet, we expect older individuals to be more prone to vaccinate because of the higher mortality risk associated with COVID-19. Also, it is unclear to what extent functional limitations and other chronic conditions have hindered some of the 80+ individuals from getting vaccinated. The consolidation of the Brazilian public health system (universal, integral, and decentralized) that resulted in expanding healthcare services and success in earlier vaccination campaigns makes us confident about the high coverage of vaccination records at older ages.

## Data Availability

We used public data only.

https://ipums.org/

https://www.ibge.gov.br/estatisticas/sociais/populacao/9109-projecao-da-populacao.html?=&t=o-que-e

https://population.un.org/wpp/

https://opendatasus.saude.gov.br/dataset/covid-19-vacinacao/resource/ef3bd0b8-b605-474b-9ae5-c97390c197a8

https://mortality.org/

https://www.gov.br/saude/pt-br/coronavirus/publicacoes-tecnicas/guias-e-planos/plano-nacional-de-vacinacao-covid-19/view

## Footnotes

3 The excluded sets intersect. Therefore, the final total number of excluded cases (missing dose, missing sex, vaccination date before January 18th, healthcares workers born in December 30, 1899) totalize only 16,006 (0.024%).

4 Japan: Statistics Bureau of Japan (2007; 2014; 2017). Sweden: Statistiska Centralbyran (2003; 2004; 2005; 2006; 2007; 2008), Statistics Sweden (2010; 2011; 2012; 2015; 2017; 2018; 2019; 2020). Switzerland: Swiss Federal Statistical Office (SFSO) (2012; 2016; 2018; 2020).

## References

Brasil. 2021a. Plano Nacional de Operacionalização da Vacinação Contra a COVID-19. 6th ed. Brasília, DF.

Brasil. 2021b. “Campanha Nacional de Vacinação contra COVID-19 - Registros de Vacinação COVID-19 - Open Data.” openDataSUS. 2021. https://opendatasus.saude.gov.br/dataset/covid-19-vacinacao/resource/ef3bd0b8-b605-474b-9ae5-c97390c197a8.

Carrier, N. H., and A. M. Farrag. 1959. “The Reduction of Errors in Census Populations for Statistically Underdeveloped Countries.” Population Studies 12 (3): 240–85. https://doi.org/10.1080/00324728.1959.10405023.

Coale, Ansley J., and Ellen Eliason Kisker. 1986. “Mortality Crossovers: Reality or Bad Data?” Population Studies 40 (3): 389–401. https://doi.org/10.1080/0032472031000142316.

Conselho Nacional de Secretários de Saúde - CONASS. 2021. “Painel de Análise do Excesso de Mortalidade por Causas Naturais No Brasil.” 2021. https://www.conass.org.br/indicadores-de-obitos-por-causas-naturais/.

Dechter, Aimée R, and Samuel H Preston. 1991. “Age Misreporting and Its Effects on Adult Mortality Estimates in Latin America.” Population Bulletin of the United Nations, no. 31–32: 1–16.

Di Lego, Vanessa, Cássio M. Turra, and Cibele Cesar. 2017. “Mortality Selection among Adults in Brazil: The Survival Advantage of Air Force Officers.” Demographic Research 37 (October): 1339–50. https://doi.org/10.4054/DemRes.2017.37.41.

Elo, Irma T., and Samuel H. Preston. 1994. “Estimating African-American Mortality from Inaccurate Data.” Demography 31 (3): 427–58. https://doi.org/10.2307/2061751.

Glei, Dana, Andres Barajas Paz, Jose Manuel Aburto, and Magali Barbieri. 2021. “Mexican Mortality 1990-2016: Comparison of Unadjusted and Adjusted Estimates.” Demographic Research 44 (April): 719–58. https://doi.org/10.4054/DemRes.2021.44.30.

Gomes, Marilia Miranda Fortes, and Cassio M. Turra. 2009. “The Number of Centenarians in Brazil: Indirect Estimates Based on Death Certificates.” Demographic Research 20 (April): 495–502. https://doi.org/10.4054/DemRes.2009.20.20.

Grushka, Carlos Oscar. 1996. “Adult and Old Age Mortality in Latin America: Evaluation, Adjustments and a Debate over a Distinct Pattern.” PhD Dissertation, University of Pennsylvania.

Hakkert, Ralph. 1996. Fontes de dados demográficos. Textos didáticos 3. Belo Horizonte: Associação Brasileira de Estudos Populacionais.

Human Mortality Database. 2021. University of California, Berkeley (USA) and Max Planck Institute for Demographic Research (Germany). www.mortality.org or www.humanmortality.de.

IBGE. 2013. Projeções Da População: Brasil e Unidades da Federação. Séries Relatórios Metodológicos, volume 40. Rio de Janeiro: IBGE, Coordenação de População e Indicadores Sociais. https://www.ibge.gov.br/estatisticas/sociais/populacao/9109-projecao-da-populacao.htm.

IBGE, 2013a. Tábuas Abreviadas de Mortalidade Por Sexo e Idade: Brasil, Grandes Regiões e Unidades da Federação, 2010. Estudos e Pesquisas. Informação Demográfica e Socioeconômica, número 30. Rio de Janeiro: IBGE, Coordenação de População e Indicadores Sociais.

IBGE. 2018. Projeções Da População: Brasil e Unidades da Federação - Revisão 2018. 2nd ed. Séries Relatórios Metodológicos, volume 40. Rio de Janeiro: IBGE, Coordenação de População e Indicadores Sociais. https://www.ibge.gov.br/estatisticas/sociais/populacao/9109-projecao-da-populacao.htm.

Lundström, H. 2002. “Annual Population Register Counts as of December 31st, 1990-2001. Unpublished Computer File.” Human Mortality DataBase RefCode 39.

Minnesota Population Center. 2020. “Integrated Public Use Microdata Series, International: Version 7.3.” Minneapolis, MN: IPUMS. https://doi.org/10.18128/D020.V7.3.

Myers, Robert J. 1954. “Accuracy of Age Reporting in the 1950 United States Census.” Journal of the American Statistical Association 49 (268): 826–31. https://doi.org/10.1080/01621459.1954.10501237.

Nepomuceno, Marília R., and Cássio M. Turra. 2020. “The Population of Centenarians in Brazil: Historical Estimates from 1900 to 2000.” Population and Development Review 46 (4): 813–33. https://doi.org/10.1111/padr.12355.

Palloni, Alberto, Hiram Beltrán-Sánchez, and Guido Pinto. 2021. “Estimation of Older-Adult Mortality from Information Distorted by Systematic Age Misreporting.” Population Studies, May, 1–18. https://doi.org/10.1080/00324728.2021.1918752.

Preston, Samuel H., Irma T. Elo, Ira Rosenwaike, and Mark Hill. 1996. “African-American Mortality at Older Ages: Results of a Matching Study.” Demography 33 (2): 193–209. https://doi.org/10.2307/2061872.

Preston, Samuel H., Irma T. Elo, and Quincy Stewart. 1999. “Effects of Age Misreporting on Mortality Estimates at Older Ages.” Population Studies 53 (2): 165–77. https://doi.org/10.1080/00324720308075.

Rosenwaike, Ira, and Samuel H. Preston. 1984. “Age Overstatement and Puerto Rican Longevity.” Human Biology 56 (3): 503–25.

Siegel, Jacob S., and David A. Swanson, eds. 2004. The Methods and Materials of Demography. Second edition. New York: Elsevier/Academic Press.

Statistics Bureau of Japan. 2007. “The 2005 Population Census of Japan, Vol. 2-1 Sex, Marital Status and Marital Status of Population.” Human Mortality DataBase RefCode 96.

Statistics Bureau of Japan. 2014. “Annual October 1st Population Estimates for Years 1996-2012. Reference Table Two. Years 2000, 2005 and 2010 Are Census Counts for Single-Ages 0-100+ (2000) and 0-115 (2005 and 2010), with Population of Unknown Age Already Redistributed. Intercensal Years Are Official Estimates for Single-Ages 0-90+. Data Were Collected and Sent as a Spreadsheet by Futoshi Ishii of the National Institute of Population and Social Security Research.” Human Mortality DataBase RefCode 103.

Statistics Bureau of Japan. 2017. “2015 Population Census. Data Sent as a Spreadsheet by Futoshi Ishii of the National Institute of Population and Social Security Research.” Human Mortality DataBase RefCode 116.

Statistics Sweden. 2010. “Swedish Population on December 31, 2008 and 2009, by Sex and Age. Retrieved from Statistics Sweden Website (http://www.scb.se). Population for Ages 100 and over Has Been Received Directly from NSO.” Human Mortality DataBase RefCode 15.

Statistics Sweden. 2011. “Swedish Population on December 31, 2010, by Sex and Age. Received from Statistics Sweden.” Human Mortality DataBase RefCode 17.

Statistics Sweden. 2012. “Swedish Population on December 31, 2011, by Sex and Age. Retrieved from the Statistics Sweden Website (http://www.scb.se).” Human Mortality DataBase RefCode 55.

Statistics Sweden. 2015. “Population by Year, Single Year of Age and Sex. 2012-2014. Sweden - Data Have Been Received Directly from Statistics Sweden.” Human Mortality DataBase RefCode 60.

Statistics Sweden. 2017. “Population by Year, Single Year of Age and Sex. 2015-2016. Sweden - Data Have Been Received Directly from Statistics Sweden.” Human Mortality DataBase RefCode 68.

Statistics Sweden. 2018. “Population by Year, Age and Sex, Single Year 2017. Sweden - Data Have Been Received Directly from Statistics Sweden.” Human Mortality DataBase RefCode 72.

Statistics Sweden. 2019. “Population by Year, Age and Sex, Single Year 2018. Sweden - Data Have Been Received Directly from Tomas Johansson at Statistics Sweden.” Human Mortality DataBase RefCode 76.

Statistics Sweden. 2020. “Population by Year, Age and Sex, Single Year 2019. Sweden - Data Have Been Received Directly from Tomas Johansson at Statistics Sweden.” Human Mortality DataBase RefCode 80.

Statistiska Centralbyran. 2003. “Tabell 1.1 - Population by Sex and Age in One-Year Groups and Five-Year Groups, in the Whole Country, Dec. 31, 2002.” Human Mortality DataBase RefCode 42.

Statistiska Centralbyran. 2004. “Tabell 1.5 - Population by Sex, Age (in One-Year and Five-Year Groups) and Marital Status on Dec. 31, 2003.” Human Mortality DataBase RefCode 45.

Statistiska Centralbyran. 2005. “Swedish Population on December 31, 2004, by Sex and Age.” Human Mortality DataBase RefCode 47.

Statistiska Centralbyran. 2006. “Swedish Population on December 31, 2005, by Sex and Age.” Human Mortality DataBase RefCode 48.

Statistiska Centralbyran. 2007. “Swedish Population on December 31, 2006, by Sex and Age.” Human Mortality DataBase RefCode 51.

Statistiska Centralbyran. 2008. “Swedish Population on December 31, 2007, by Sex and Age.” Human Mortality DataBase RefCode 53.

Swiss Federal Statistical Office (SFSO). 2012. “December 31st Population Estimates, 2001-2009. Ages 0-98 from SFSO. Complemented with January 1st Eurostat Population Estimates for Ages 99-110+ (Years 2002-2010).” Human Mortality DataBase RefCode 27.

Swiss Federal Statistical Office (SFSO). 2016. “Population, 2010-2014. Received as an Electronic File in an Email Attachment from Corinne Di Loreto.” Human Mortality DataBase RefCode 31.

Swiss Federal Statistical Office (SFSO). 2018. “December 31st Population, 2010-2016. Received as an Electronic File in an Email Attachment from Dominik Ullmann.” Human Mortality DataBase RefCode 35.

Swiss Federal Statistical Office (SFSO). 2020. “January 1st Population, 2018-2020. Received as an Electronic File in an Email Attachment from Corinne Di Loreto.” Human Mortality DataBase RefCode 39.

Turra, Cássio M. 2012. “Os Limites Do Corpo: A Longevidade Em Uma Perspectiva Demográfica.” Revista Da Universidade Federal de Minas Gerais 19 (1 e 2): 156–81.

United Nations. 2019. World Population Prospects 2019: Online Edition. New York: United Nations, Department of Economic and Social Affairs, Population Division. https://population.un.org/wpp/.

